# Effects of Buprenorphine, Methadone and Substance-Use on COVID-19 Morbidity and Mortality

**DOI:** 10.1101/2023.11.24.23298995

**Authors:** Nicholaus Christian, Xin Zhou, Rajiv Radhakrishnan

## Abstract

**Objectives:** Substance use disorder has been associated with increased morbidity in COVID-19 infection. However, less is known about the impact of active substance use and medications for opioid use disorder (MOUD) on COVID-19 outcomes. We conducted a retrospective cohort study to evaluate the impact of substance use, namely cannabis, cocaine, alcohol, sedative and opioid use as well as buprenorphine or methadone = on COVID-19 morbidity and mortality.

**Methods:** Using electronic-health record data at a large urban hospital system, patients who tested positive for COVID-19 between January 1, 2020 to December 31, 2021 were included. Substance use was identified from urine toxicology and MOUD prescriptions within 90 days prior to admission. COVID-19 outcomes included mortality, ICU admission, need for ventilatory support, number and duration of hospitalizations. Multivariable logistic regression was performed controlling for variables such as age, sex, medical comorbidity, tobacco use, and social disadvantage.

**Results:** Among COVID-19 positive patients (n=17,423), sedative, cannabis, cocaine, and opioid use was associated with statistically significant increases in need for ICU care, need for ventilatory support, number of hospitalizations and duration of hospitalization. Substance use was not associated with an increase in all-cause mortality. There were no statistically significant differences between methadone, buprenorphine and other opioids on COVID-19 outcomes.

**Conclusions:** Active substance use were associated with increased morbidity in COVID-19 infection. MOUD was not associated with worse COVID-19 outcomes compared to OUD. Future studies focused on MOUD treatments that reduce morbidity may help improve clinical outcomes in COVID-19.

## INTRODUCTION

People who use substances are disproportionately impacted by the novel SARS-CoV-2 virus and associated coronavirus disease (COVID-19) given social marginalization, distrust of the healthcare system, and predisposition to poor respiratory health.^1,2^ Studies investigating COVID-19 outcomes for people who use substances have generally demonstrated an increase in hospitalizations and worsening of COVID-19 severity,^3,4^ although data on mortality have been conflicting. Multiple studies have shown that the increase in mortality from COVID-19 among people with a substance use disorder (SUD) diagnosis disappears after accounting for physical health comorbidities.^5,6^ This suggests that the diagnosis of SUD alone is not what is responsible for the worse COVID-19 outcomes— it is the physical health comorbidities that develop as a consequence of long term substance use. Additionally, medications for opioid use disorder (MOUD) such as buprenorphine and methadone have not been shown to increase the risk of contracting COVID-19 or cause worsening of COVID-19 outcomes.^7,8^ However, no study to date has compared the COVID-19 outcomes of individuals on MOUD versus non-prescribed opioids.

Previous studies have also focused on the diagnosis of SUD in the electronic medical record as opposed to investigating if current substance use might impact COVID-19 outcomes. Due to this, it is unclear if substance use alone is the causal factor of the worse outcomes or if it is the social factors and health comorbidities that accompany a history of chronic substance use that is driving these differences. An alternative exposure variable to indicate active substance use is urine toxicology screening, although to date no studies have investigated if positivity on urine toxicology screening is associated with COVID-19 morbidity and mortality.

## METHODS

### Study Procedures

We conducted a retrospective cohort study of all patients with the diagnosis of COVID-19 that were admitted to a large urban health care system in New Haven, Connecticut between January 2020 and December 2021. Data were extracted from the electronic health records (EHR). The study was approved for exemption of review and waiver of consent by the institutional review board of Yale University. This report followed the Strengthening the Reporting of Observational Studies in Epidemiology ([STROBE](https://www.equator-network.org/wp-content/uploads/2015/10/STROBE_checklist_v4_cohort.pdf)) reporting guideline for cohort studies.^9^

### Measures

The exposure variables of interest were urine drug screening (UDS) positivity for benzodiazepines/barbiturates (sedatives), cannabis, cocaine and opioids, and documented outpatient prescription of buprenorphine or methadone within 90 days prior to admission. The outcome variables of interest a priori were mortality, ICU admission, length of ICU admission, need for ventilatory support, days on ventilatory support, number of hospital admissions, and total length of hospitalization.

### Statistical Analysis

Multivariable linear regression and multivariable logistic regression were performed controlling for age, sex, ethnicity, race, disability, medical comorbidity (Charlson Comorbidity Index), tobacco use, social disadvantage (derived from zip code), and psychiatric diagnosis. Missing values on patient characteristics were handled by multiple imputation ^10^.

## RESULTS

### Patient Characteristics

17,423 patients were admitted for COVID-19 infection during the study period, (53.9% female, 57.5% were white, and 41.8% were older than 65.) Rates of sedative, cannabis, cocaine, and opioid positivity were 1.5%, 2.0%, 1.4%, and 3.4% respectively, with 53 patients taking buprenorphine and 216 taking methadone at admission (see Table 1).

**Table 1.** Demographic characteristics, substance use, and COVID-related outcomes.

| **Characteristics** | **Cohort (n=17,423)** |
| --- | --- |
| **Age** |  |
| 18-64, n (%) | 10,134 (58.2) |
| 65+, n (%) | 7,289 (41.8) |
| **Sex** |  |
| Female, n (%) | 9,395 (53.9) |
| Male, n (%) | 8,028 (46.1) |
| **Ethnicity** |  |
| Hispanic or Latino, n (%) | 3,990 (22.9) |
| Non-Hispanic, n (%) | 13,271 (76.2) |
| **Race** |  |
| White or Caucasian, n (%) | 10,013 (57.5) |
| Black or African American, n (%) | 3,732 (21.4) |
| Other, n (%) | 3,441 (19.8) |
| **Disabled** |  |
| Yes, n (%) | 1,679 (9.6) |
| No, n (%) | 15,663 (89.9) |
| **Tobacco Use** |  |
| Yes, n (%) | 1,658 (9.5) |
| Quit, n (%) | 6,159 (35.4) |
| Never, n (%) | 8,728 (50.1) |
| **Charlson Comorbidity Index** | 1.93 ± 2.24 |
| **Social disadvantage** | 0.12 ± 0.08 |
| **Psychiatric diagnosis, n (%)** | 1,158 (6.7) |
| **Substance use** |  |
| Sedatives, n (%) | 268 (1.5) |
| Cannabinoid, n (%) | 350 (2.0) |
| Cocaine, n (%) | 236 (1.4) |
| Opioids, n (%) | 585 (3.4) |
| Methadone, n (%) | 216 (1.2) |
| Buprenorphine, n (%) | 53 (0.3) |
| Methadone / buprenorphine, n (%) | 259 (1.5) |
| **Outcomes** |  |
| All-cause mortality, n (%) | 2,260 (13.0) |
| # hospitalizations | 1.6 ± 1.4 |
| Total duration of hospitalization (days) | 14.1 ± 22.7 |
| In ICU ever? n (%) | 4,303 (24.7) |
| Duration in ICU (days) | 2.6 ± 11.1 |
| On Vent ever? n (%) | 2,073 (11.9) |
| Duration on vent (days) | 1.2 ± 6.5 |

### Association Between Substance Use and COVID-19 Outcomes

Substance use was not associated with an increase in all-cause mortality compared to patients without substance use (OR 0.82, 95% CI 0.65 to 1.06; P =.14). However, increases in duration of hospitalization were noted with sedatives (12.67, 95% CI 10.11 to 15.22; P<.01), cannabinoids (4.76, 95% CI 2.50 to 7.03; P<.01), cocaine (9.90, 95% CI 7.12 to 12.67; P<.01), and opioids (12.26, 95% CI 10.47 to 14.05; P<.01) compared to a non-substance using cohort. In addition, substance use was associated with increases in number of hospitalizations and likelihood of intubation. Sedative and opioid use were associated with increased likelihood of ICU admissions, and increased ICU length of stay.

### Association Between Medications for Opioid Use Disorder and COVID-19 Outcomes

After adjusting socioeconomic and other variables, no statistically significant difference was found in the tested COVID outcomes between methadone, buprenorphine, prescribed vs non-prescribed opioids.

## DISCUSSION

In this retrospective cohort study using EHR data, we found that sedatives (benzodiazepine/barbiturates), cannabis, cocaine, and opioid UDS positivity was associated with increased COVID-19 morbidity in terms of need for ICU care, need for ventilatory support, number of hospitalizations and duration of hospitalization. The association between UDS positivity and increased hospital length of stay for individuals with COVID-19 affirms previous studies concluding that a diagnosis of SUD was associated with prolonged hospitalization.^4,8^ These findings are consistent with our earlier study (Ramakrishnan et al.^4^) and the study by Vai et al, that found that OUD was associated with prolonged hospitalization compared to the general population even prior to COVID-19.^11^ Opioid and sedative positivity was associated with longer critical care length of stay and ventilator use. This may be explained by the effects of these substances on respiratory depression.^8^ The impact of cannabis use on COVID-19 morbidity is particularly important in view of the fact that medical cannabis dispensaries were allowed to operate as an “essential” business during the COVID-19 pandemic while other businesses were required to shut down in 2020. The wave of legalization of medical cannabis and recreational cannabis across states in the US has also resulted in decreased perception of harm from cannabis^12,13^.

We did not find an association between sedatives, cannabis, cocaine, and opioid use (including MOUD) and COVID-19 mortality after adjusting for covariates including medical comorbidity. This is consistent with other studies that show that although SUD is associated with worsening of COVID-19 morbidity, the trend towards worse mortality disappears after accounting for comorbidities.^4-6,14^ Only one study has noted a 3% increase in mortality that remained after adjusting for medical comorbidity, however, they were unable to examine mortality for SUD subtypes due to small sample sizes in the study.^15^ These findings suggest that SUDs might only be a predictor for mortality inasmuch as chronic substance use might cause comorbid respiratory and cardiovascular diseases. In a national case-control study, among individuals with SUD, those with diabetes or chronic lung disease were more likely to present to the hospital for COVID-19.^16^

The similarity in outcomes between methadone, buprenorphine, and non-prescribed opioids may be explained by common pharmacological effects of opioid-related respiratory depression, complex opioid immune modulation, and drug-drug interactions associated with opioid use.^17^ People on buprenorphine and methadone were also noted to have higher rates of hospitalization compared to individuals without OUD in a prior study.^8^ This has important implications for clinicians counseling patients with SUDs regarding COVID-19 risk factors for severe disease. MOUD are not more dangerous than non-prescribed opioids, and therefore should be continued during illness given the benefit of reduction of overdose mortality. Additionally, patients should be counseled on possible worse COVID-19 outcomes if actively using substances, thus patients hoping to reduce their risk of severe COVID-19 should be counseled to discontinue using substances, especially if feeling ill.

There are important limitations to this study. The first is that although UDS testing gives a snapshot of someone’s use patterns, it does not delineate use severity or confer a diagnosis of substance use disorder. Secondly, this was a single site study at an institution that had good COVID-19 outcomes which might skew the results. Third, at least 8 months of the study period occurred prior to the development of the COVID-19 vaccine. The associations found in this study might be an overestimation of actual risk given that the vaccine is so protective against severe COVID-19.

## CONCLUSIONS

In this retrospective cohort study, substance use was associated with worse COVID-19 morbidity, but not mortality. Medications for OUD were found to have the similar COVID-19 outcomes as non-prescribed opioids. Thus, the benefits of treating OUD with methadone/buprenorphine (and consequently reducing the risk of overdose) greatly outweigh the risks of severe COVID-19.

## Data Availability

All data produced in the present study are available upon reasonable request to the authors

## ACKNOWLEDGEMENTS

Yale Center for Clinical Investigations (YCCI) Joint Data Analytics Team (JDAT).

## 2. Analysis results

|  | Mortality |  | # of hospital admissions |  | Total length of hospitalization |  | ICU admission |  | Length of ICU admission |  | Need for ventilatory support |  | Days on ventilatory support |  |
| --- | --- | --- | --- | --- | --- | --- | --- | --- | --- | --- | --- | --- | --- | --- |
|  | Odds ratio (95% CI) | P | Difference (95% CI) | P | Difference (95% CI) | P | Odds ratio (95% CI) | P | Difference (95% CI) | P | Odds ratio (95% CI) | P | Difference (95% CI) | P |
| All substance use * | 0.82<br>(0.64, 1.06) | 0.14 | 0.85<br>(0.76, 0.93) | <0.01 | 9.0<br>(7.58, 10.44) | <0.01 | 1.52<br>(1.30, 1.78) | <0.01 | 0.74<br>(-0.01, 1.50) | 0.05 | 1.81<br>(1.51, 2.18) | <0.01 | 0.22<br>(-0.22, 0.66) | 0.33 |
| Sedatives * | 0.88<br>(0.56, 1.39) | 0.59 | 1.35<br>(1.20, 1.51) | <0.01 | 12.67<br>(10.11, 15.22) | <0.01 | 2.16<br>(1.66, 2.82) | <0.01 | 1.39<br>(0.04, 2.74) | 0.04 | 2.62<br>(1.94, 3.52) | <0.01 | 0.34<br>(-0.45, 1.12) | 0.40 |
| Cannabinoid * | 0.72<br>(0.44, 1.19) | 0.20 | 0.60<br>(0.46, 0.75) | <0.01 | 4.76<br>(2.50, 7.03) | <0.01 | 1.24<br>(0.95, 1.60) | 0.11 | 0.06<br>(-1.14, 1.25) | 0.93 | 1.57<br>(1.16, 2.14) | 0.004 | -0.28<br>(-0.97, 0.42) | 0.44 |
| Cocaine * | 0.83<br>(0.48, 1.42) | 0.49 | 0.74<br>(0.57, 0.91) | <0.01 | 9.90<br>(7.12, 12.67) | <0.01 | 1.20<br>(0.88, 1.64) | 0.25 | 0.16<br>(-1.30, 1.62) | 0.83 | 1.42<br>(0.99, 2.04) | 0.06 | 0.29<br>(-0.57, 1.14) | 0.51 |
| Opioids * | 0.94<br>(0.70, 1.27) | 0.70 | 1.04<br>(0.93, 1.15) | <0.01 | 12.26<br>(10.47, 14.05) | <0.01 | 1.44<br>(1.19, 1.74) | <0.01 | 0.95<br>(0.00, 1.89) | 0.05 | 1.81<br>(1.45, 2.26) | <0.01 | 0.43<br>(-0.12, 0.98) | 0.13 |
| prescribed vs. non-prescribed opioids † | 0.79<br>(0.43, 1.43) | 0.43 | -0.12<br>(-0.67, 0.43) | 0.66 | 2.88<br>(-3.91, 9.68) | 0.41 | 0.84<br>(0.56, 1.24) | 0.37 | 0.94<br>(-0.88, 2.76) | 0.31 | 0.91<br>(0.59, 1.40) | 0.66 | 1.23<br>(-0.36, 2.83) | 0.13 |

\* The sample size was 17,423.
† The analysis (methadone/buprenorphine vs. non-prescribed opioids) was performed within the subset of subjects who took opioids, and the sample size was 585,

## REFERENCES

1. Becker WC, Fiellin DA. When Epidemics Collide: Coronavirus Disease 2019 (COVID-19) and the Opioid Crisis. Ann Intern Med. Jul 7 2020;173(1):59–60. doi:10.7326/M20-1210

2. Volkow ND. Collision of the COVID-19 and Addiction Epidemics. Ann Intern Med. Jul 7 2020;173(1):61–62. doi:10.7326/M20-1212

3. Schieber LZ, Dunphy C, Schieber RA, Lopes-Cardozo B, Moonesinghe R, Guy GP, Jr. Hospitalization Associated With Comorbid Psychiatric and Substance Use Disorders Among Adults With COVID-19 Treated in US Emergency Departments From April 2020 to August 2021. JAMA Psychiatry. Feb 15 2023;doi:10.1001/jamapsychiatry.2022.5047

4. Ramakrishnan D, Sureshanand S, Pittman B, Radhakrishnan R. Impact of Cannabis Use, Substance Use Disorders, and Psychiatric Diagnoses on COVID-19 Outcomes: A Retrospective Cohort Study. J Clin Psychiatry. Aug 29 2022;83(5)doi:10.4088/JCP.21m14332

5. Allen B, El Shahawy O, Rogers ES, Hochman S, Khan MR, Krawczyk N. Association of substance use disorders and drug overdose with adverse COVID-19 outcomes in New York City: January-October 2020. J Public Health (Oxf). Sep 22 2021;43(3):462–465. doi:10.1093/pubmed/fdaa241

6. Baillargeon J, Polychronopoulou E, Kuo YF, Raji MA. The Impact of Substance Use Disorder on COVID-19 Outcomes. Psychiatr Serv. May 1 2021;72(5):578–581. doi:10.1176/appi.ps.202000534

7. Owiti JA, Benson M, Maplanka M, Oluseye L, Carvalho D. Is Methadone Safe for Patients With Opioid Use Disorder and Coronavirus Disease 2019 Infection? J Addict Nurs. Apr-Jun 01 2022;33(2):86–94. doi:10.1097/JAN.0000000000000457

8. Qeadan F, Tingey B, Bern R, et al. Opioid use disorder and health service utilization among COVID-19 patients in the US: A nationwide cohort from the Cerner Real-World Data. EClinicalMedicine. Jul 2021;37:100938. doi:10.1016/j.eclinm.2021.100938

9. von Elm E, Altman DG, Egger M, et al. The Strengthening the Reporting of Observational Studies in Epidemiology (STROBE) statement: guidelines for reporting observational studies. Bull World Health Organ. Nov 2007;85(11):867–72. doi:10.2471/blt.07.045120

10. Little RJR, D.B. Statistical Analysis with Missing Data. vol 793. John Wiley & Sons; 2019.

11. Alemu BT, Olayinka O, Martin BC. Characteristics of Hospitalized Adults with Opioid Use Disorder in the United States: Nationwide Inpatient Sample. Pain Physician. Aug 2021;24(5):327–334.

12. Syed SA, Singh J, Elkholy H, et al. Impact of personal beliefs about medical cannabis on physician recommendation practices: Results of an international survey. Asian J Psychiatr. Jul 2023;85:103634. doi:10.1016/j.ajp.2023.103634

13. Harris-Lane LM, Storey DP, Drakes DH, Donnan JR, Bishop LD, Harris N. Emerging adult perceptions of cannabis consumption: Examining changes in perceptions from pre-legalization to post-legalization. Int J Drug Policy. Oct 2023;120:104193. doi:10.1016/j.drugpo.2023.104193

14. Vai B, Mazza MG, Delli Colli C, et al. Mental disorders and risk of COVID-19-related mortality, hospitalisation, and intensive care unit admission: a systematic review and meta-analysis. Lancet Psychiatry. Sep 2021;8(9):797–812. doi:10.1016/S2215-0366(21)00232-7

15. Wang QQ, Kaelber DC, Xu R, Volkow ND. COVID-19 risk and outcomes in patients with substance use disorders: analyses from electronic health records in the United States. Mol Psychiatry. Jan 2021;26(1):30–39. doi:10.1038/s41380-020-00880-7

16. Board AR, Kim S, Park J, et al. Risk factors for COVID-19 among persons with substance use disorder (PWSUD) with hospital visits - United States, April 2020-December 2020. Drug Alcohol Depend. Mar 1 2022;232:109297. doi:10.1016/j.drugalcdep.2022.109297

17. Schimmel J, Manini AF. Opioid Use Disorder and COVID-19: Biological Plausibility for Worsened Outcomes. Subst Use Misuse. 2020;55(11):1900–1901. doi:10.1080/10826084.2020.1791184

